# Molecular alterations in TP53, WNT, PI3K, TGF-Beta and RTK/RAS pathways in gastric cancer among ethnically heterogeneous cohorts

**DOI:** 10.1101/2025.02.22.25322719

**Authors:** Cecilia Monge, Brigette Waldrup, Francisco G. Carranza, Enrique Velazquez-Villarreal

## Abstract

**Background/Objectives:** Gastric cancer (GC) remains a leading cause of cancer-related mortality worldwide, with significant racial and ethnic disparities in incidence, molecular characteristics, and patient outcomes. However, genomic studies focusing on Hispanic/Latino (H/L) populations remain scarce, limiting our understanding of ethnicity-specific molecular alterations. This study aims to characterize pathway-specific mutations in TP53, WNT, PI3K, TGF-Beta and RTK/RAS signaling pathways in GC and compare mutation frequencies between H/L and Non-Hispanic White (NHW) patients. Additionally, we evaluate the impact of these alterations on overall survival using publicly available datasets.

**Methods:** We conducted a bioinformatics analysis using publicly available GC datasets to assess mutation frequencies in TP53, WNT, PI3K, TGF-Beta and RTK/RAS pathway genes. A total of 800 patients were included in the analysis, comprising 83 H/L patients and 717 NHW patients. Patients were stratified by ethnicity (H/L vs. NHW) to evaluate differences in mutation prevalence. Chi-squared tests were performed to compare mutation rates between groups, and Kaplan-Meier survival analysis was used to assess overall survival differences based on pathway alterations among both H/L and NHW patients.

**Results:** Significant differences were observed in the TP53 pathway and related genes when comparing GC in H/L patients to NHW patients. TP53 mutations were less prevalent in H/L patients (9.6% vs. 19%, p = 0.03). Borderline significant differences were noted in the WNT pathway when comparing GC in H/L patients to NHW GC patients, with WNT alterations more frequent in H/L GC (8.4% vs. 4%, p = 0.08), and APC mutations significantly higher (3.6% vs. 0.8%, p = 0.05). Although alterations in PI3K, TGF-Beta and RTK/RAS pathways were not statistically significant, borderline significance was observed in genes related to these pathways, including EGFR (p = 0.07), FGFR1 (p = 0.05), FGFR2 (p = 0.05), and PTPN11 (p = 0.05) in the PI3K pathway, and SMAD4 (p = 0.08) in the TGF-Beta pathway. Survival analysis revealed no significant differences among H/L patients. However, NHW patients with TP53 and PI3K pathway alterations exhibited significant differences in overall survival, while those without TGF-Beta pathway alterations also showed a significant survival impact. In contrast, WNT pathway alterations were not associated with significant survival differences. These findings suggest that TP53, PI3K, and TGF-Beta pathway disruptions may have distinct prognostic implications in NHW GC patients.

**Conclusions:** This study provides one of the first ethnicity-focused analyses of TP53, WNT, PI3K, TGF-Beta and RTK/RAS pathway alterations in GC, revealing significant racial/ethnic differences in pathway dysregulation. The findings suggest that TP53 and WNT alterations may play a critical role in GC among H/L patients, while PI3K and TGF-Beta alterations may have greater prognostic significance in NHW patients. These insights emphasize the need for precision medicine approaches that account for genetic heterogeneity and ethnicity-specific pathway alterations to improve cancer care and outcomes for underrepresented populations.

## 1. Introduction

Gastric cancer (GC) remains one of the most prevalent and lethal malignancies worldwide, ranking as the fourth leading cause of cancer-related deaths globally (1,2). Although advances in diagnostic tools and treatment strategies have improved survival rates in high-income countries, GC continues to disproportionately impact specific racial and ethnic populations (3). Notably, Hispanic/Latino (H/L) individuals face a higher incidence of GC and worse survival outcomes compared to Non-Hispanic White (NHW) patients (4–7). These disparities persist even after adjusting for socioeconomic factors, suggesting underlying biological differences that warrant further investigation (8–10). While previous studies have explored the molecular landscape of GC in Asian and NHW populations, genomic alterations specific to the H/L population remain largely underexplored.

Molecular profiling studies have identified five key signaling pathways involved in GC development and progression: TP53, WNT, PI3K, TGF-Beta, and RTK/RAS. These pathways regulate essential cellular functions such as proliferation, apoptosis, DNA repair, and immune response, and their dysregulation is frequently associated with aggressive tumor behavior, therapy resistance, and poor prognosis (11–13). Understanding pathway-specific alterations in H/L GC patients is crucial for uncovering ethnicity-specific molecular drivers that contribute to disease progression and disparities in clinical outcomes.

The TP53 signaling pathway plays a pivotal role in genomic stability, apoptosis, and cell cycle regulation (14). Mutations in TP53 are among the most common alterations in GC, with studies reporting mutation rates as high as 30-50% (15). Helicobacter pylori (H. pylori) infection, a well-established risk factor for GC, has been implicated in TP53 mutations through genomic instability and DNA damage (16). Furthermore, TP53 alterations have been linked to chemoresistance, potentially limiting treatment efficacy in GC patients (17). However, the prevalence and prognostic impact of TP53 mutations in H/L GC patients remain poorly characterized.

The WNT signaling pathway, particularly the WNT/β-catenin axis, is a key regulator of cell proliferation and differentiation. Aberrant activation of this pathway has been observed in intestinal-type GC, where mutations in genes such as RNF43 (3-44%) and LRP1B (31-67%) drive tumor progression (18). Studies suggest that WNT ligand overexpression in GC promotes cancer stem cell renewal and enhances invasive properties (19). Given the high rates of late-stage diagnosis among H/L GC patients, it is essential to determine whether WNT pathway alterations contribute to more aggressive disease phenotypes in this population.

The PI3K/AKT/mTOR signaling pathway is a major driver of tumor metabolism, growth, and survival. PI3K pathway dysregulation, often through PIK3CA mutations and PTEN deletions, is associated with increased tumor invasiveness, immune evasion, and resistance to therapy (20). In GC, PI3K/AKT pathway hyperactivation has been linked to poor prognosis and reduced response to chemotherapy (21). Moreover, H. pylori infection has been shown to activate PI3K/AKT signaling, further underscoring its role in GC pathogenesis (22). While PI3K/AKT pathway alterations have been extensively studied in other populations, their impact on GC outcomes in H/L patients remains unclear.

The TGF-Beta signaling pathway plays a paradoxical role in GC, functioning as both a tumor suppressor in cancer and a pro-oncogenic driver in advanced disease (23). In later stages, TGF-Beta promotes epithelial-to-mesenchymal transition (EMT), angiogenesis, and immune suppression, leading to increased tumor progression and metastasis (24). Elevated TGF-Beta1 expression has been correlated with poor survival in GC patients (25). Additionally, H. pylori infection has been shown to activate the TGF-Beta pathway, contributing to tumor progression (26). However, it is unknown whether TGF-Beta pathway alterations contribute to disparities in GC outcomes among H/L patients.

The RTK/RAS signaling pathway regulates key cellular processes, including growth, migration, and differentiation. Mutations in KRAS, NRAS, and BRAF are frequently observed in GC, particularly in intestinal-type tumors and metastatic patients (27). These mutations drive constitutive activation of MAPK and PI3K/AKT pathways, promoting tumor progression and drug resistance (28). KRAS mutations have also been associated with resistance to EGFR-targeted therapies, which may have implications for treatment response (29) in H/L GC patients. However, the role of RTK/RAS pathway alterations in shaping the molecular landscape of H/L GC remains largely unstudied.

Given the increasing burden of GC in H/L populations and the limited molecular characterization of this disease in this group (20–34), this study aims to comprehensively analyze pathway-specific alterations in TP53, WNT, PI3K, TGF-Beta, and RTK/RAS signaling in GC. We compare mutation frequencies between H/L and NHW patients and evaluate the prognostic impact of these alterations on overall survival. By integrating genomic and survival data, this study seeks to provide novel insights into ethnicity-specific molecular differences that may contribute to GC disparities. These findings may inform precision medicine strategies and guide targeted therapeutic interventions to improve GC outcomes in underrepresented populations.

## 2. Materials and Methods

To conduct our analysis, we utilized clinical and genomic data from 11 GC datasets accessed via the cBioPortal database. These datasets included studies classified under GC, as well as data from the GENIE BPC GC v2.0-public dataset. Two studies focusing on metastatic GC were excluded to ensure the analysis was limited to primary tumor patients. Following dataset selection, we applied a series of filtering criteria to refine our sample pool. Patients were included if they were identified as H/L. This process resulted in three datasets meeting all criteria comprising 83 H/L GC patients. For NHW patients, 717 GC patients were included using the same inclusion criteria but applied within this specific racial and ethnic group (Tables 1 & 2). This study represents one of the largest comprehensive characterizations of TP53, WNT, PI3K, TGF-Beta and RTK/RAS pathway alterations in an underserved population, providing critical insights into the molecular disparities in GC.

Ethnicity-based classification stratified participants into H/L and NHW groups. We further stratified these groups based on the presence or absence of TP53, WNT, PI3K, TGF-Beta and RTK/RAS pathway alterations, enabling a detailed examination of the interactions between ethnicity, and these molecular changes. Table 1 presents the number of patients included in the analysis of H/L and NHW patients, with a total of 83 H/L patients and 717 NHW patients. This analysis evaluates the prevalence of TP53, WNT, PI3K, TGF-Beta and RTK/RAS pathway alterations by comparing H/L and NHW GC patients. By integrating these stratifications, our study provides one of the most comprehensive characterizations of TP53, WNT, PI3K, TGF-Beta and RTK/RAS pathway disruptions in an underserved population, offering valuable insights into potential molecular disparities and their implications for precision medicine in GC.

**Table 1.** Patient Demographics and Clinical Characteristics of the Hispanic/Latino (H/L) and non-Hispanic White (NHW) gastric cancer (GC) cohorts.

| Clinical Feature | H/L Cohort<br>n (%) | NHW Cohort<br>n (%) |
| --- | --- | --- |
| **Gender** |  |  |
| Male | 38 (45.8%) | 387 (54.0%) |
| Female | 45 (54.2%) | 330 (46.0%) |
| **Tumor Type** |  |  |
| Primary | 83 (100.0%) | 717 (100.0%) |
| Metastatic | 0 (0.0%) | 0 (0.0%) |
| **Ethnicity** |  |  |
| Spanish/Hispanic/Latino | 83 (100.0%) | 0 (0.0%) |
| Non-Spanish; Non-Hispanic | 0 (0.0%) | 717 (100.0%) |

Statistical analysis included Chi-square tests to evaluate the independence of categorical variables and identify potential associations between ethnicity, and pathway alterations. Additionally, we stratified samples by tumor location, distinguishing between colon and rectal cancers. This level of stratification facilitated a nuanced analysis of the interplay between ethnicity, and tumor location in relation to pathway disruptions, offering deeper insights into patient heterogeneity and potential implications for treatment responses. Kaplan-Meier survival analysis was employed to assess overall survival, focusing on the impact of TP53, WNT, PI3K, TGF-Beta and RTK/RAS pathway alterations. Survival curves were constructed to illustrate survival probabilities over time, with patients grouped by the presence or absence of pathway disruptions. The log-rank test was utilized to determine statistically significant differences between survival curves. Median survival times were calculated, accompanied by 95% confidence intervals to convey the precision of these estimates. This comprehensive methodological approach provided an in-depth understanding of how specific pathway alterations may affect patient outcomes in GC patients within the H/L population.

## 3. Results

From the cBioPortal projects mentioned above that reported ethnicity, we identified and constructed our H/L cohort, which comprised 83 samples, while the NHW cohort included 717 samples (Table 1). The H/L cohort consisted of 45.8% male and 54.2% female patients, whereas the NHW cohort comprised 54% male and 46% female patients. At the time of diagnosis, all patients presented with primary tumors. Additionally, all patients in the H/L cohort self-identified as Hispanic or Latino, ensuring the study’s focus on this targeted demographic.

The comparative analysis of genomic features between H/L and NHW patients, reveals notable distinctions (Table 2). The median mutation count was lower in the H/L cohort (2 mutations, IQR: 1-2) compared to the NHW cohort (2 mutations, IQR: 1-4), with a p-value of 0.01266, indicating a statistically significant difference. Similarly, the median TMB was lower in H/L patients (0.865 mutations/Mb, IQR: 0.865-1.73) compared to NHW patients (1.6 mutations/Mb, IQR: 0.9-2.2), with a highly significant p-value of 0.00175. These findings suggest that H/L patients exhibit a lower overall mutational burden in comparison to NHW patients. Additionally, the median FGA, which represents the fraction of the genome affected by copy number alterations, was lower in H/L patients (0.101, IQR: 0.03-0.15) than in NHW patients (0.142, IQR: 0.06-0.26), with a p-value of 0.002095. This suggests that H/L GC patients may have fewer structural alterations and chromosomal instability compared to NHW patients. Overall, these results highlight significant genomic differences between H/L and NHW GC patients, with H/L patients exhibiting lower mutation counts, TMB, and FGA, which may have implications for tumor biology and therapeutic responses in these populations.

**Table 2.** Ethnicity-associated differences in clinical features between Hispanic/Latino (H/L) and non-Hispanic White (NHW) gastric cancer (GC) cohorts.

| Clinical Feature | H/L Samples<br>n (%) | NHW Samples<br>n (%) | p-value |
| --- | --- | --- | --- |
| Median Mutation Count (IQR)* | 2 (1-2) | 2 (1-4) | **0.01266** |
| Median TMB (IQR)** | 0.865 (0.865-1.73) | 1.6 (0.9-2.2) | **0.00175** |
| Median FGA (IQR)*** | 0.101 (0.03-0.15) | 0.142 (0.06-0.26) | **0.002095** |
| **Oncotree Code** |  |  |  |
| GIST | 83 (100.0%) | 717 (100.0%) | NA |
| **APC Mutation** |  |  |  |
| Present | 3 (3.6%) | 6 (0.8%) | 0.05708 |
| Absent | 80 (96.4%) | 711 (99.2%) |  |
| **SMAD4 Mutation** |  |  |  |
| Present | 2 (2.4%) | 3 (0.4%) | 0.08645 |
| Absent | 81 (97.6%) | 714 (99.6%) |  |
| **EGFR Mutation** |  |  |  |
| Present | 3 (3.6%) | 7 (1.0%) | 0.07558 |
| Absent | 80 (96.4%) | 710 (99.0%) |  |
| **FGFR1 Mutation** |  |  |  |
| Present | 3 (3.6%) | 6 (0.8%) | 0.05708 |
| Absent | 80 (96.4%) | 711 (99.2%) |  |
| **FGFR2 Mutation** |  |  |  |
| Present | 2 (2.4%) | 2 (0.3%) | 0.05557 |
| Absent | 81 (97.6%) | 715 (99.7%) |  |
| **PTPN11 Mutation** |  |  |  |
| Present | 2 (2.4%) | 2 (0.3%) | 0.05557 |
| Absent | 81 (97.6%) | 715 (99.7%) |  |

\*HL NA: 5, NHW NA: 48
\*\*HL NA: 66, NHW NA: 421
\*\*\*HL NA: 40, NHW NA: 391

**Table 3.** Rates of TP53, WNT, PI3K, TGF-Beta and RTK/RAS pathway alterations among Hispanic/Latino (H/L) and Non-Hispanic White (NHW) gastric cancer (GC) patients.

|  | H/L Samples<br>n (%) | NHW Samples<br>n (%) | p-value |
| --- | --- | --- | --- |
| TP53 Alterations Present | 8 (9.6%) | 136 (19.0%) | **0.03489** |
| TP53 Alterations Absent | 75 (90.4%) | 581 (81.0%) |  |
| WNT Alterations Present | 7 (8.4%) | 29 (4.0%) | 0.08674 |
| WNT Alterations Absent | 76 (91.6%) | 688 (96.0%) |  |
| PI3K Alterations Present | 8 (9.6%) | 108 (15.1%) | 0.2477 |
| PI3K Alterations Absent | 75 (90.4%) | 609 (84.9%) |  |
| TGF-beta Alterations Present | 2 (2.4%) | 10 (1.4%) | 0.3586 |
| TGF-beta Alterations Absent | 81 (97.6%) | 707 (98.6%) |  |
| RTK/RAS Alterations Present | 74 (89.2%) | 635 (88.6%) | 1 |
| RTK/RAS Alterations Absent | 9 (10.8%) | 82 (11.4%) |  |

Furthermore, within the WNT pathway, APC mutations were found to be significantly more prevalent in H/L GC patients compared to NHW patients (3.6% vs. 0.8%, p = 0.05). While no overall statistical significance was observed in the PI3K, TGF-Beta, and RTK/RAS pathways, several genes exhibited borderline significance, suggesting potential ethnic-specific variations in GC molecular profiles. In the PI3K pathway, alterations in EGFR (p = 0.07), FGFR1 (p = 0.05), FGFR2 (p = 0.05), and PTPN11 (p = 0.05) were observed, while SMAD4 (p = 0.08) exhibited borderline significance in the TGF-Beta pathway. These findings underscore the importance of ethnicity-specific molecular characterization in GC and may provide insights into pathogenesis and potential therapeutic targets for underrepresented populations.

In our analysis of genetic alterations in GC among H/L and NHW individuals, we observed notable differences in the frequency of TP53 and WNT mutations, with TP53 alterations reaching statistical significance. TP53 mutations were present in 9.6% of H/L patients, compared to 19.0% of NHW patients (p = 0.03489), indicating a significantly lower prevalence of TP53 alterations in the H/L cohort. Conversely, the absence of TP53 mutations was more frequent in H/L patients (90.4%) compared to NHW patients (81.0%), further highlighting this disparity. Although WNT pathway alterations were not statistically significant, they were observed at a higher frequency in H/L patients (8.4%) compared to NHW patients (4.0%, p = 0.08674), suggesting a possible trend that warrants further investigation. PI3K pathway alterations were found in 9.6% of H/L patients and 15.1% of NHW patients (p = 0.2477), indicating no significant difference between the groups. Similarly, TGF-Beta pathway alterations were present in 2.4% of H/L patients and 1.4% of NHW patients (p = 0.3586), with no substantial disparity observed. Interestingly, RTK/RAS pathway alterations were highly prevalent in both cohorts, with 89.2% of H/L patients and 88.6% of NHW patients exhibiting these mutations (p = 1), suggesting a shared molecular signature in GC development across both ethnic groups. These findings highlight potential differences in the genetic landscape of GC between H/L and NHW patients, particularly with lower TP53 mutation prevalence in H/L individuals, which may have implications for tumor biology, disease progression, and targeted treatment approaches. Further research is necessary to explore the functional impact of these mutations and their potential role in ethnic-specific GC disparities.

The Kaplan-Meier survival analysis for H/L GC patients with TP53 pathway alterations indicated no statistically significant difference in overall survival between those with and without the alteration (Figure 1). A total of 14 patients were in the altered group, while 69 patients were in the not altered group, with identical survival trajectories observed in both groups (p = 1). The overlapping confidence intervals highlight high variability in the survival estimates at different time points, suggesting that TP53 pathway alterations may not be a strong prognostic factor in this cohort. However, the small sample size may limit statistical power, and further studies with larger datasets are needed to better assess the potential impact of TP53 alterations on GC outcomes.

**Figure 1.**
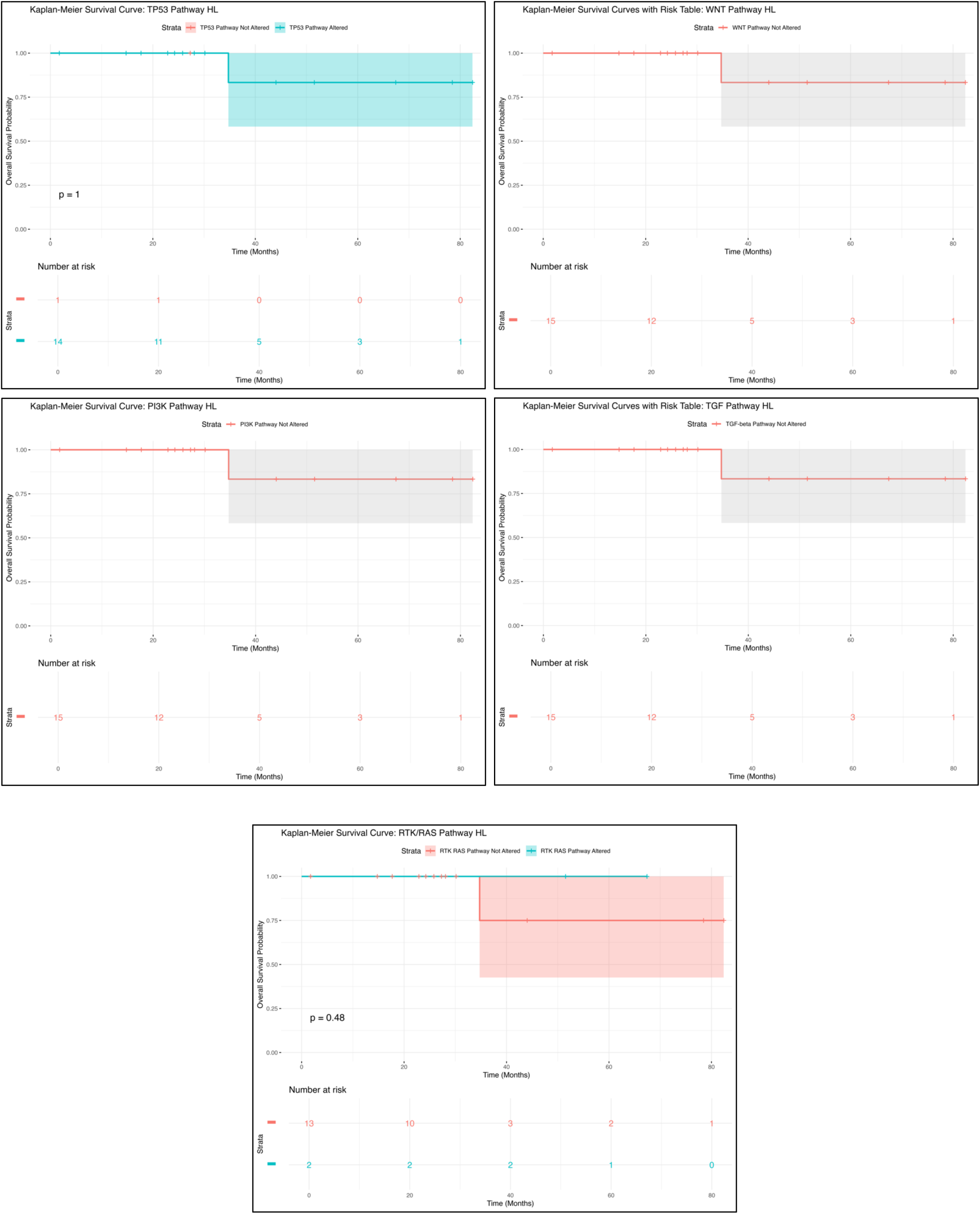
Kaplan-Meier overall survival curves for Hispanic/Latino (H/L) gastric cancer (GC) patients, stratified by the presence or absence of TP53 (upper left), WNT (upper right), PI3K (middle left), TGF-Beta (middle right), and RTK/RAS (lower center) pathway alterations.

Similarly, the Kaplan-Meier survival analysis for WNT pathway alterations showed no statistically significant difference in survival outcomes (Figure 1). Among the 19 patients with WNT pathway alterations, the survival curve remained relatively stable, with a single drop in survival occurring at approximately 30 months, followed by a plateau. The wide confidence intervals surrounding the survival curve suggest a high degree of uncertainty, likely due to the limited sample size. These findings indicate that WNT pathway alterations may not have a strong prognostic impact, though further validation with larger datasets is necessary to confirm these observations.

For PI3K pathway alterations, the Kaplan-Meier survival analysis revealed no statistically significant survival differences (Figure 1). A total of 19 patients were in the PI3K pathway altered group, but no separate comparison group was available. The survival curve exhibited a drop in survival at approximately 30 months, followed by a plateau, with wide confidence intervals suggesting considerable variability. These findings indicate that PI3K pathway alterations may not serve as a strong prognostic marker in this cohort, though further investigation is warranted to assess their potential role in GC progression.

Similarly, the TGF-Beta pathway alterations displayed a comparable trend to the PI3K pathway (Figure 1). Among the 19 patients with TGF-Beta alterations, the survival curve showed a decline at approximately 30 months, followed by a plateau, with high variability due to the small sample size. No statistically significant survival differences were observed, reinforcing the need for larger studies to validate the potential prognostic implications of TGF-Beta pathway alterations in GC.

Lastly, the Kaplan-Meier survival analysis for RTK/RAS pathway alterations (Figure 1) showed no significant difference in survival between patients with and without alterations (p = 0.48). Of the patients analyzed, 7 were in the altered group, while 12 were in the not altered group. The survival trajectories between both groups were nearly identical, and the overlapping confidence intervals suggest high uncertainty in survival estimates. Given the small sample size, additional research with larger cohorts is necessary to further explore the prognostic relevance of RTK/RAS pathway alterations in GC.

Overall, these findings emphasize the uncertainty in survival differences across multiple pathway alterations, underscoring the need for larger and more diverse datasets to better understand the molecular drivers of GC in H/L populations and their potential implications for targeted therapies.

The Kaplan-Meier survival analysis for TP53 pathway alterations in NHW GC patients indicated no statistically significant difference in overall survival between those with and without the alteration (Figure S1). A total of 14 patients were in the altered group, while 69 patients were in the not altered group, and the survival trajectories of both groups appeared nearly identical (p = 1). The overlapping confidence intervals and minimal divergence in survival curves suggest that TP53 pathway alterations may not be a strong prognostic factor in NHW GC patients. However, given the small sample size of the altered group, statistical power may be limited, potentially obscuring subtle survival differences. Further research with larger patient cohorts is necessary to clarify the role of TP53 alterations in this population.

In contrast, WNT pathway alterations in NHW GC patients showed a slight trend toward reduced survival compared to those without alterations (Figure S1). Among the 46 patients in the altered group and 460 patients in the not altered group, the survival curve of WNT-altered patients (red curve) exhibited a modest decline in survival probability relative to the non-altered group (blue curve). However, this difference was not statistically significant (p = 0.6), and the wide confidence intervals suggest a high degree of uncertainty, likely due to the smaller sample size in the altered group. While these findings do not provide strong evidence of WNT alterations affecting survival outcomes, further stratification or larger datasets may be necessary to assess their clinical relevance more comprehensively.

The Kaplan-Meier survival analysis for PI3K pathway alterations in NHW GC patients revealed a statistically significant association with poorer overall survival (Figure S1). Among the 97 patients in the altered group and 602 in the not altered group, patients with PI3K pathway alterations (red curve) had markedly worse survival outcomes compared to those without alterations (blue curve) (p < 0.0001). The clear separation between the survival curves supports the notion that PI3K pathway alterations may serve as a negative prognostic factor in NHW GC. The wider confidence intervals in the altered group suggest some variability in survival estimates, but the statistical significance underscores the potential impact of PI3K dysregulation on disease progression. These findings warrant further investigation into the clinical implications of PI3K alterations, particularly in the context of targeted therapeutic approaches.

Similarly, TGF-Beta pathway alterations were associated with significantly worse survival outcomes in NHW GC patients (Figure S1). Among the 56 patients with TGF-Beta pathway alterations and 486 in the not altered group, patients harboring TGF-Beta pathway mutations (red curve) demonstrated a noticeable decline in survival probability compared to those without alterations (blue curve). This difference was statistically significant (p = 0.016), suggesting that TGF-Beta pathway dysregulation may contribute to disease progression and worse prognosis. However, the wide confidence intervals around the altered group’s survival curve indicate variability in survival estimates, likely due to the small sample size. Further research is needed to assess the biological mechanisms underlying these alterations and their potential as therapeutic targets in NHW GC.

In contrast, RTK/RAS pathway alterations did not show a statistically significant impact on overall survival in NHW GC patients (Figure S1). Among the 57 patients in the altered group and 517 in the not altered group, the survival trajectories of both groups were nearly identical, with no significant difference observed (p = 0.99). The overlapping survival curves and wide confidence intervals suggest that RTK/RAS pathway alterations may not play a major prognostic role in this cohort. However, given the small number of altered cases, additional studies with larger datasets may help determine whether RTK/RAS alterations influence survival outcomes in specific GC subtypes.

Overall, these results highlight pathway-specific differences in survival outcomes among NHW GC patients, with PI3K and TGF-Beta pathway alterations showing significant associations with poorer prognosis, whereas TP53, WNT, and RTK/RAS alterations did not appear to strongly influence survival outcomes. These findings underscore the importance of genomic profiling in NHW GC patients and suggest that targeting PI3K and TGF-Beta pathways may offer potential therapeutic benefits. Further studies with larger and more diverse patient populations are needed to better understand the clinical implications of these molecular alterations and their potential role in precision medicine strategies for GC treatment.

The alteration rates of TP53, WNT, PI3K, TGF-Beta, and RTK/RAS pathway-related genes were analyzed among H/L and NHW GC patients to determine potential ethnicity-related differences (Table S1). The analysis revealed that most genes associated with the TP53 pathway, including MDM2, MDM4, RPS6KA3, and CHEK2, displayed low alteration rates in both H/L and NHW patients, with no statistically significant differences. TP53 mutations were slightly more prevalent in H/L patients (6.0%) compared to NHW patients (4.5%), but this difference was not statistically significant (p = 0.5765). Similarly, alterations in ATM and CDKN2A were observed at low frequencies in both groups, with no significant differences detected. These findings suggest that TP53 pathway mutations are relatively rare in both populations, and their role in GC progression may be influenced by additional molecular or environmental factors.

Within the WNT pathway, APC mutations were found to be more frequent in H/L GC patients (3.6%) compared to NHW patients (0.8%), showing a borderline statistical significance (p = 0.05708). Other WNT pathway-related genes, including CTNNB1, AXIN2, and TCF7L2, exhibited low alteration rates with no significant differences between the two groups. These findings suggest that while APC mutations may be more common in H/L patients, further investigation is needed to determine their potential impact on tumor biology and disease progression.

For the PI3K pathway, PIK3CA and PTEN mutations were observed at higher frequencies in H/L patients (3.6%) compared to NHW patients (1.8%), but these differences were not statistically significant (p = 0.2257). Other PI3K-related genes, including PIK3R1, PIK3R2, AKT1, and MTOR, had low or absent mutation rates in both groups, with no significant differences. These findings suggest that PI3K pathway alterations may occur at comparable rates across ethnic groups, with no clear evidence of ethnicity-specific molecular drivers in this pathway.

In the TGF-Beta pathway, SMAD4 mutations were more prevalent in H/L patients (2.4%) compared to NHW patients (0.4%), with borderline statistical significance (p = 0.08645). However, other genes in this pathway, including TGFBR1, TGFBR2, SMAD2, and SMAD3, exhibited low or absent mutation rates in both groups, with no significant differences. The trend toward a higher frequency of SMAD4 mutations in H/L patients suggests a possible role in GC development, but larger datasets are needed to confirm this observation and determine its clinical significance.

Lastly, analysis of the RTK/RAS pathway revealed that EGFR mutations were more frequent in H/L patients (3.6%) compared to NHW patients (1.0%), showing a borderline significance (p = 0.07558). Similarly, FGFR1 and FGFR2 mutations were more prevalent in H/L patients (3.6% and 2.4%, respectively) compared to NHW patients (0.8% and 0.3%, respectively), both showing borderline significance (p = 0.05708 and p = 0.05557, respectively). However, KRAS, NRAS, BRAF, and MET alterations did not show significant differences between the two groups, suggesting that mutations in key oncogenic drivers are largely similar across ethnicities.

These findings highlight potential ethnicity-specific trends in GC molecular alterations, particularly in APC, SMAD4, EGFR, and FGFR1/2 genes. However, the lack of statistical significance in most comparisons underscores the need for larger studies to validate these observations and explore their implications for precision medicine and targeted therapy in H/L GC patients.

## 4. Discussion

GC remains a leading cause of cancer-related mortality worldwide, with significant racial and ethnic disparities in both incidence and outcomes. While extensive molecular profiling has been conducted in Asian and NHW populations, data on the genomic landscape of GC in H/L patients remain limited. This study aimed to characterize the molecular alterations in TP53, WNT, PI3K, TGF-Beta, and RTK/RAS pathways in GC and explore their potential prognostic implications by comparing H/L and NHW patients. Our findings revealed ethnicity-specific differences in key molecular alterations, particularly within the TP53 and WNT pathways, with potential clinical implications for risk stratification and precision medicine approaches.

Our analysis revealed a significantly lower prevalence of TP53 mutations in H/L patients (9.6%) compared to NHW patients (19%, p = 0.03). TP53 plays a crucial role in maintaining genomic stability and preventing tumor progression. The lower mutation frequency in H/L patients suggests potential differences in the mechanisms of tumor development between ethnic groups. Since TP53 alterations are frequently associated with poor prognosis, genomic instability, and therapy resistance, their reduced occurrence in H/L GC patients raises questions about alternative pathways driving tumorigenesis in this population. It is also possible that epigenetic modifications, rather than direct TP53 mutations, contribute to GC progression in H/L individuals, warranting further investigation through multi-omics analyses.

The WNT pathway alterations were more common in H/L patients (8.4%) than in NHW patients (4%, p = 0.08), suggesting a possible trend toward increased WNT pathway dysregulation in the H/L population. Specifically, APC mutations were significantly more prevalent in H/L patients (3.6% vs. 0.8%, p = 0.05), which may have implications for disease progression and tumor behavior. The WNT/β-catenin signaling pathway plays a central role in GC pathogenesis by regulating cell proliferation, differentiation, and stemness. The increased prevalence of APC mutations in H/L patients suggests that WNT pathway activation may serve as a distinct molecular driver in this population, which could influence tumor aggressiveness and treatment response. Previous studies have shown that WNT pathway activation correlates with increased cancer stem cell properties and chemoresistance, highlighting the potential need for targeted therapies aimed at this pathway in H/L GC patients.

Although alterations in the PI3K, TGF-Beta, and RTK/RAS pathways did not reach statistical significance, several genes within these pathways exhibited borderline significance, hinting at possible ethnicity-related differences in mutation profiles. Notably, within the PI3K pathway, genes such as EGFR (p = 0.07), FGFR1 (p = 0.05), FGFR2 (p = 0.05), and PTPN11 (p = 0.05) exhibited marginally higher alteration rates in H/L patients compared to NHW patients. These findings suggest that PI3K pathway dysregulation may still play a role in H/L GC, particularly through receptor tyrosine kinase (RTK) activation, even if the overall pathway alteration rate was not significantly different. Additionally, SMAD4 alterations in the TGF-Beta pathway (p = 0.08) were slightly more frequent in H/L patients, pointing to a potential role for TGF-Beta signaling in GC pathogenesis in this population. Given the dual role of TGF-Beta as both a tumor suppressor and a pro-oncogenic factor, further studies are needed to explore whether SMAD4 alterations contribute to differential disease progression between ethnic groups.

Kaplan-Meier survival analysis revealed ethnicity-specific differences in the prognostic impact of these pathway alterations. Among NHW GC patients, PI3K and TGF-Beta pathway alterations were significantly associated with worse overall survival (p < 0.0001 and p = 0.016, respectively), whereas no significant survival differences were observed for TP53, WNT, or RTK/RAS alterations. In contrast, H/L GC patients showed no significant survival differences based on pathway alterations. These findings suggest that PI3K and TGF-Beta pathway alterations may have a greater prognostic impact in NHW GC patients than in H/L patients. The reasons for this discrepancy remain unclear but may involve differences in tumor microenvironment, immune response, or genetic ancestry. Additionally, the smaller sample size of H/L patients may have limited statistical power, underscoring the need for larger, well-powered studies to further investigate these potential prognostic differences.

The significant association between PI3K pathway alterations and poorer survival in NHW patients supports previous studies demonstrating that PI3K/AKT/mTOR signaling is a critical driver of tumor progression and therapy resistance in GC. Given that PI3K pathway alterations have been linked to resistance to EGFR-targeted therapies, our findings suggest that NHW GC patients with PI3K alterations may benefit from alternative therapeutic strategies targeting this pathway, such as PI3K inhibitors or combination therapies. Conversely, the lack of a significant survival impact in H/L patients suggests that additional molecular mechanisms may contribute to GC progression in this group, reinforcing the need for ethnicity-specific research.

Similarly, TGF-Beta pathway alterations were significantly associated with worse survival in NHW patients but not in H/L patients. This aligns with previous studies indicating that high TGF-Beta1 expression is linked to lower survival rates in GC patients. The TGF-Beta signaling pathway is known to promote epithelial-to-mesenchymal transition (EMT), immune evasion, and metastasis in advanced GC. The observed survival disparity raises intriguing questions about whether TGF-Beta pathway activation exerts distinct biological effects in H/L and NHW patients, potentially due to differences in immune response or tumor microenvironment (31).

Despite the strengths of this study, including one of the largest ethnicity-specific analyses of pathway alterations in GC, several limitations must be acknowledged. The retrospective nature of publicly available datasets introduces potential biases, including differences in sample collection, sequencing depth, and patient demographics. Furthermore, the underrepresentation of H/L patients in genomic databases remains a major challenge in cancer research. Our findings highlight the urgent need for larger, prospective studies focusing on H/L populations to fully characterize GC molecular disparities. Additionally, functional validation studies are required to determine whether the observed pathway alterations translate into meaningful biological differences in tumor progression and therapy response.

## 5. Conclusions

In conclusion, this study provides important insights into the molecular landscape of GC in H/L patients, revealing significant differences in TP53 and WNT pathway alterations compared to NHW patients. While TP53 mutations were significantly less frequent in H/L patients, APC mutations were more common, suggesting potential ethnicity-specific drivers of tumor progression. Additionally, PI3K and TGF-Beta pathway alterations were significantly associated with poor survival in NHW patients, but not in H/L patients, highlighting possible differences in the prognostic impact of these pathways across ethnic groups. These findings emphasize the importance of ethnicity-specific genomic research to better understand the biological underpinnings of GC disparities and inform the development of precision medicine strategies tailored to underrepresented populations. Future studies integrating multi-omics analyses, immune profiling, and clinical outcomes data will be essential to uncover the full extent of molecular differences and improve therapeutic interventions for H/L GC patients.

## Data Availability

All data used in the present study is publicly available at https://www.cbioportal.org/ and https://genie.cbioportal.org. Additional data can be provided upon reasonable request to the authors.

## Notes

### Competing Interest Statement

The authors have declared no competing interest.

### Funding Statement

This study was funded by the National Cancer Institute, NCI, award number U2CCA252971; the City of Hope Cancer Control and Population Sciences program by the National Institutes of Health, NIH, National Cancer Institute, NCI, award number P30CA033572; and the Drug Development and Capacity Building: A UCR/CoH-CCC Partnership project by the National Institutes of Health, NIH, National Cancer Institute, NCI, award number U54 CA285116.

### Author Declarations

The source data used in this study were publicly available before the initiation of the study and can be accessed through cBioPortal for Cancer Genomics at https://www.cbioportal.org/ and the GENIE Project (AACR Project GENIE cBioPortal) at https://genie.cbioportal.org. Additional data may be provided upon reasonable request to the authors

